# Effectiveness of BNT162b2 and CoronaVac against pediatric COVID-19-associated hospitalization and moderate-to-severe disease

**DOI:** 10.1101/2022.09.09.22279426

**Authors:** Jaime S. Rosa Duque, Daniel Leung, Ka Man Yip, Derek H.L. Lee, Hung-kwan So, Wilfred H.S. Wong, Yu Lung Lau

**Affiliations:** Department of Paediatrics and Adolescent Medicine, The University of Hong Kong, Hong Kong, China

**Keywords:** COVID-19, vaccine, BNT162b2, CoronaVac, Omicron, adolescents, children

## Abstract

**Background:** Vaccine effectiveness (VE) of BNT162b2 and CoronaVac against COVID-19-associated hospitalization and moderate-to-severe disease due to SARS-CoV-2 Omicron BA.2 for pediatric populations that had low exposure to prior SARS-CoV-2 variants needs to be further clarified. This can be studied from the 1.36 million vaccine doses had been administered to 766,601 of 953,400 children and adolescents in Hong Kong (HK) since March 2021 to April 2022.

**Methods:** Using an ecological design leveraging the HK vaccination coverage statistics and public hospital records, this study investigated the VE for children aged 3-11 years and adolescents aged 12-18 years at the population level during the Omicron BA.2 wave from January to April 2022.

**Findings:** VE against COVID-19-associated hospitalization for children was 65.3% for 1 dose of BNT162b2 and 13.0% and 86.1% for 1 and 2 doses of CoronaVac, respectively. For adolescents, VE against COVID-19-associated hospitalization was 60.2% and 82.4% after 1 and 2 doses of BNT162b2 and 30.8% and 90.7% after 1 and 2 doses of CoronaVac, respectively. Protection against moderate-to-severe disease for aged 3-18 was high, with VE of 93.1% and 95.8% after 2 doses of BNT162b2 and CoronaVac, respectively. No COVID-19-associated hospitalization or moderate-to-severe disease occurred for 68,565 children and adolescents who received their third dose. Estimated hospitalizations of children and adolescents averted by vaccination were 68 and 999, respectively, and were 45 and 147 for moderate-to-severe cases.

**Conclusions:** BNT162b2 or CoronaVac provide substantial protection from COVID-19-associated hospitalization and moderate-to-severe disease due to a SARS-CoV-2 variant of concern.

**Funding:** The Providence Foundation.

## INTRODUCTION

The SARS-CoV-2 Omicron variant continues to cause millions of COVID-19-associated pediatric hospitalizations, severe disease and death globally.^1-4^ BNT162b2 and CoronaVac are amongst the top most widely used COVID-19 vaccines across the world.^5^ In Hong Kong (HK), these two vaccines have been authorized for adolescents since 16 Mar 2021 and then children since 21 Jan 2022, while the third dose of both vaccines for all healthy adolescents and children commenced on 11 Mar 2022 and 14 Apr 2022, respectively (Figure S1). Consequently, 1.36 million vaccine doses had been administered to 766,601 (80.4%) of the total HK pediatric population of 953,400 from March 2021 to April 2022, with 331,948 (65.6%) of 506,100 who were children aged 3-11 years (aged 5-11 for BNT162b2 and 3-11 years for CoronaVac) and 434,653 (97.1%) of 447,300 who were adolescents aged 12-18 years.^6^ Of 258,466 children, 73,482 had received at least one dose of BNT162b2 (22.1% amongst those vaccinated) or CoronaVac (77.9%), respectively, while 174,152 (34.4% of the total HK pediatric population) remained unvaccinated (Figure S2). For adolescents, 372,753 and 61,900 had received at least one dose of BNT162b2 (85.8%) or CoronaVac (14.2%), respectively, whereas 12,627 (2.8%) remained unvaccinated.

The early phases of such a widespread vaccination program relied on data from licensing trials that demonstrated non-inferior neutralization and efficacy against COVID-19 for BNT162b2 in those aged 5-17 and immunogenicity for CoronaVac in aged 3-17 but not effectiveness against hospitalization or severe disease, despite these endpoints are the most relevant real-life outcomes.^7-9^ Evaluation of real-life vaccine effectiveness (VE) against hospitalization and severe disease in children and adolescents is important since they have different immune responses to vaccines than adults.^10^ VE studies performed in children and adolescents during Omicron BA.1 waves revealed lower VE against symptomatic COVID-19 and generally preserved VE against severe outcomes,^11-22^ yet isolated studies showed low VE against hospitalization or severe outcomes.^12,23^ Moreover, pediatric VE data on the subvariant BA.2, which is antigenically distinct from BA.1, are lacking.^24,25^ A major Omicron wave predominated by the highly transmissible BA.2 in HK caused many pediatric hospitalizations, clinical complications and deaths in early 2022 despite the continuation of the elimination strategy that had suppressed SARS-CoV-2 circulation in the past.^1,6^

During this BA.2 wave, essentially all hospitalized pediatric COVID-19 cases received medical care in public hospitals, all of which utilize the same electronic health software operated by the HK Hospital Authority (HA). The HK Department of Health (DH) had instituted a rigorous tracking system on COVID-19 vaccinations and compulsory reporting of infections. These recent events and the established infrastructure that maintains such robust archive of data have created the possibility to study VE against severe disease by BA.2 in a population unexposed to prior SARS-CoV-2 variants.^26,27^

By retrieving population-level vaccine coverage statistics from DH and clinical data of hospitalized pediatric patients with laboratory confirmed COVID-19 from HA, we aimed to determine the VE against COVID-19-associated hospitalization and moderate-to-severe disease by BNT162b2 in children and adolescents aged 5-18 years and CoronaVac in children and adolescents aged 3-18 years during the January 2022 to April 2022 BA.2 wave using an ecological study design. In addition, based on the VE results, we investigated the whole population impact of vaccination, with estimations of cases of COVID-19-associated hospitalization and moderate-to-disease averted by BNT162b2 and CoronaVac in children and adolescents.

## RESULTS

During the study period of January 2022 (21 Jan 2022 for children aged 3-11 years and 1 Jan 2022 for adolescents aged 12-18 years) to 19 Apr 2022, there were 1,099 and 455 COVID-19-associated hospitalizations for children and adolescents, respectively (Table 1). Over this study period, the daily numbers of COVID-19-associated hospitalization were relatively higher for children (Figure 1A) than adolescents (Figure 1B), which peaked in late January to February 2022 for both age groups. A similar pattern was observed for the daily numbers of COVID-19-associated moderate-to-severe disease, but the peak occurred later, in late February 2022 (Figure 1C and Figure 1D). 197 children and 31 adolescents had COVID-19-associated moderate-to-severe disease (Table 1). There were few patients with comorbidities (Table S1).

**Table 1.** Numbers for COVID-19-associated hospitalization and moderate-to-severe disease for children (aged 3-11 years, 21 Jan 2022 to 19 Apr 2022) and adolescents (aged 12-18 years, 1 Jan 2022 to 19 Apr 2022) in the entire Hong Kong pediatric population.

|  |  | 3-11 years old |  |  |  | 12-18 years old |  |  |  |
| --- | --- | --- | --- | --- | --- | --- | --- | --- | --- |
|  |  | Female |  | Male |  | Female |  | Male |  |
|  |  | N | CHR | N | CHR | N | CHR | N | CHR |
| **Total hospitalizations** |  | 470 | 1.7% | 629 | 2.0% | 183 | 0.9% | 272 | 1.2% |
| **Vaccine types** | **Dose** |  |  |  |  |  |  |  |  |
| **BNT162b2** | 1 | 6 | 1.0% | 6 | 0.9% | 49 | 1.0% | 65 | 1.1% |
|  | 2 | N/A |  | N/A |  | 73 | 0.8% | 92 | 1.0% |
|  | 3 | N/A |  | N/A |  | N/A |  | N/A |  |
| **CoronaVac** | 1 | 53 | 1.1% | 74 | 1.3% | 7 | 0.7% | 15 | 1.4% |
|  | 2 | 2 | 0.8% | 2 | 0.7% | 2 | 0.2% | 3 | 0.3% |
|  | 3 | N/A |  | N/A |  | N/A |  | N/A |  |
| **Unvaccinated** | 0 | 409 | 1.8% | 547 | 2.1% | 52 | 1.9% | 97 | 2.6% |
|  |  | N | % | N | % | N | % | N | % |
| **Total moderate-to-severe disease** |  | 70 | 0.3% | 127 | 0.4% | 13 | 0.1% | 18 | 0.1% |
| **Vaccine types** | **Dose** |  |  |  |  |  |  |  |  |
| **BNT162b2** | 1 | 0 | 0.0% | 1 | 0.2% | 2 | 0.04% | 5 | 0.1% |
|  | 2 | N/A |  | N/A |  | 2 | 0.02% | 6 | 0.1% |
|  | 3 | N/A |  | N/A |  | 0 | 0.0% | 0 | 0.0% |
| **CoronaVac** | 1 | 14 | 0.3% | 18 | 0.3% | 0 | 0.0% | 1 | 0.1% |
|  | 2 | 1 | 0.4% | 0 | 0.0% | 0 | 0.0% | 0 | 0.0% |
|  | 3 | N/A |  | N/A |  | 0 | 0.0% | 0 | 0.0% |
| **Unvaccinated** | 0 | 55 | 0.2% | 108 | 0.4% | 9 | 0.3% | 6 | 0.2% |

N, numbers of cases; CI, confidence interval; CHR, case hospitalization ratio and % relative to published infection data<sup>6</sup>; N/A, not applicable as vaccination dose for this age group not yet commenced

**Figure 1.**
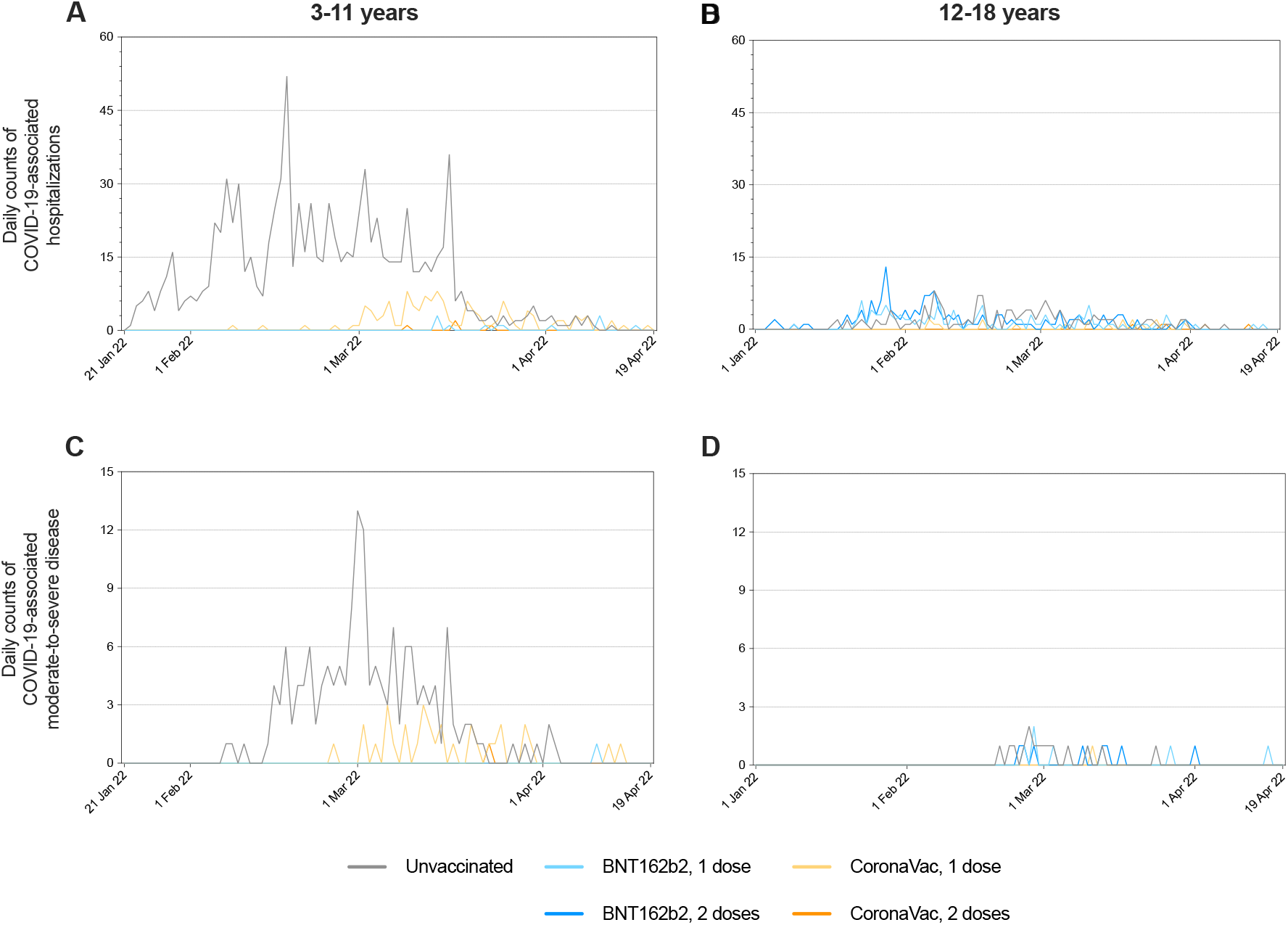
Daily numbers of COVID-19-associated hospitalizations and moderate-to-severe disease in Hong Kong by age groups and vaccine types during the study period. (A and B) The daily numbers of hospitalizations during the study period were relatively higher for children aged 3-11 years (A) (particularly the grey line, or unvaccinated) than adolescents aged 12-18 years (B). (C-D) This same pattern was observed for the daily numbers of COVID-19-associated moderate-to-severe disease, which were more for children aged 3-11 years (C) (particularly the grey line, or unvaccinated) than adolescents aged 12-18 years (D).

For VE against COVID-19-associated hospitalization, we found some protection from 1 dose of BNT162b2 in children (65.6%, 95%CI 38.2-82.5, *P*=0.0008) and adolescents (60.2%, 95%CI 47.0-70.2, *P*<0.0001) (Table 2). Two doses of CoronaVac for both children (86.2%, 95%CI 65.8-95.9, *P*=0.0001) and adolescents (90.7%, 95%CI 79.2-96.8, *P*<0.0001) and 2 doses of BNT162b for adolescents (82.3%, 95%CI 76.9-86.4, *P*<0.0001) conferred high protection. VE was unable to be estimated for 2 doses of BNT162b2 in children because the second dose in this population began at the end of the study period (BNT162b2 began on 16 Feb 2022 in children and the second dose was recommended for 8 weeks later, which was 13 Apr 2022). There was no COVID-19-associated hospitalization or moderate-to-severe disease for any of those who received 3 doses of BNT162b2 (2 children and 61,237 adolescents) or CoronaVac (40 children and 7,286 adolescents). For COVID-19-associated moderate-to-severe disease, 1 dose of BNT162b2 (84.6%, 95%CI 69.7-93.2, *P*<0.0001) and 2 doses of either BNT162b2 (93.1%, 95%CI 86.4-97.0, *P*<0.0001) or CoronaVac (95.8%, 95%CI 80.7-99.8, *P*=0.0017) conferred high protection. Results from the sensitivity analyses were similar (Table S2; Table S3).

**Table 2.** Vaccine effectiveness against COVID-19-associated hospitalization and moderate-to-severe disease with adjustment by calendar days and 14-day lag in children (aged 3-11 years, 21 Jan 2022 to 19 Apr 2022) and adolescents (aged 12-18 years, 1 Jan 2022 to 19 Apr 2022).

|  | Numbers of cases | Mean days elapsed since the last dose (SD) | VE estimate | 95%CI | p-value |
| --- | --- | --- | --- | --- | --- |
| **VE against hospitalization** |  |  |  |  |  |
| **Children (aged 3-11 years)** |  |  |  |  |  |
| *BNT162b2, 1 dose* | 12 | 27.4 (14.7) | 65.6% | 38.2 – 82.5 | 0.0008 |
| *CoronaVac, 1 dose* | 127 | 23.8 (7.5) | 13.5% | -14.0 – 34.6 | 0.3187 |
| *CoronaVac, 2 doses* | 4 | 23.3 (10.7) | 86.2% | 65.8 – 95.9 | 0.0001 |
| *Unvaccinated* | 956 | *ref* | *ref* | *ref* | *ref* |
| **Adolescents (aged 12-18 years)** |  |  |  |  |  |
| *BNT162b2, 1 dose* | 114 | 132.2 (54.1) | 60.2% | 47.0 – 70.2 | <0.0001 |
| *CoronaVac, 1 dose* | 22 | 34.7 (16.0) | 31.0% | -9.34 – 58.3 | 0.1286 |
| *BNT162b2, 2 doses* | 165 | 171.5 (51.2) | 82.3% | 76.9 – 86.4 | <0.0001 |
| *CoronaVac, 2 doses* | 5 | 47.4 (19.6) | 90.7% | 79.2 – 96.8 | <0.0001 |
| *Unvaccinated* | 149 | *ref* | *ref* | *ref* | *ref* |
| **VE against moderate-to-severe disease** |  |  |  |  |  |
| **Children and adolescents (aged 3-18 years)** |  |  |  |  |  |
| *BNT162b2, 1 dose* | 8 | 126.1 (54.0) | 84.6% | 69.7 – 93.2 | <0.0001 |
| *CoronaVac, 1 dose* | 33 | 22.2 (5.7) | 33.7% | -4.6 – 58.9 | 0.0855 |
| *BNT162b2, 2 doses* | 8 | 185.6 (49.4) | 93.1% | 86.4 – 97.0 | <0.0001 |
| *CoronaVac, 2 doses* | 1 | 39.0 (0.0) | 95.8% | 80.7 – 99.8 | 0.0017 |
| *Unvaccinated* | 178 | *ref* | *ref* | *ref* | *ref* |
| SD, standard deviation; VE, vaccine effectiveness; CI, confidence interval; ref, reference |  |  |  |  |  |

The observed (actual) and expected numbers of COVID-19-associated hospitalization in the absence of vaccination were 1,099 and 1,167 (95%CI 1,111-1,240) for children and 455 and 1,454 (95%CI 908-2,111) for adolescents, respectively. Hence, 68 (95%CI 12-141) and 999 (95%CI 453-1,656) cases of COVID-19-associated hospitalization were averted due to vaccination for children (see Methods; Figure 2A) and adolescents (Figure 2B), respectively. There were 197 and 31 cases of COVID-19-associated moderate-to-severe disease for children and adolescents, while the expected numbers were 242 (95%CI 205-244) and 178 (95%CI 93-384), respectively. The numbers of moderate-to-severe cases averted due to vaccination were 45 (95%CI 8-47) for children (Figure 2C) and 147 (95%CI 62-353) for adolescents (Figure 2D). The adjusted estimations of cases averted are depicted in Figure S3 (see Methods).

**Figure 2.**
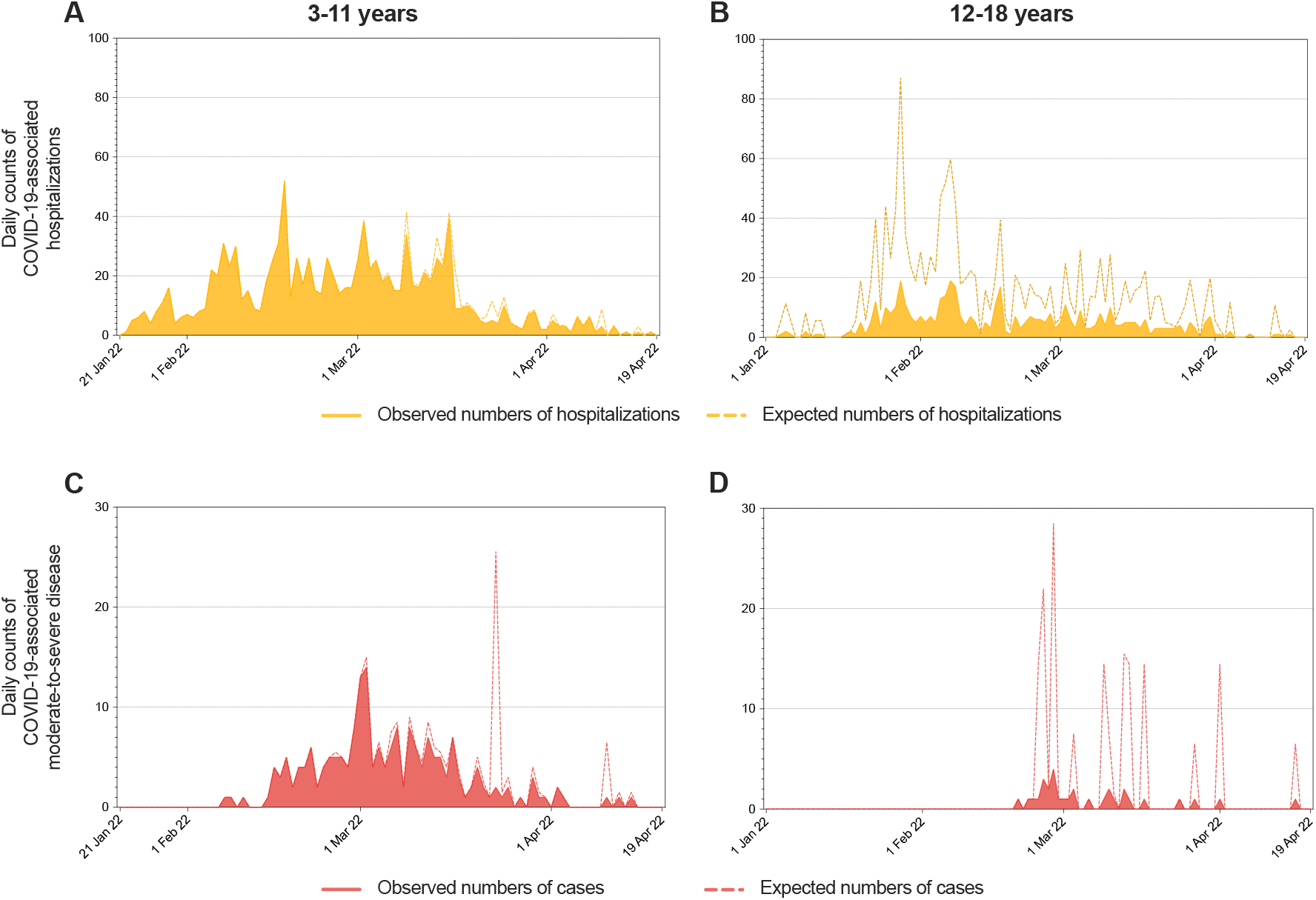
Daily numbers of COVID-19-associated hospitalizations and moderate-to-severe disease in Hong Kong averted by vaccination for children (aged 3-11 years, 21 Jan 2022 to 19 Apr 2022) and adolescents (aged 12-18 years, 1 Jan 2022 to 19 Apr 2022) (A and B) Daily numbers of expected (dotted curve) and observed (actual, orange areas) COVID-19-associated hospitalizations, with the white areas in between as the numbers of cases averted. (C and D) Daily numbers of expected (dotted curve) and observed (actual, red areas) COVID-19 associated moderate-to-severe disease, with the white areas in between as the numbers of cases averted.

## DISCUSSION

While VE had been shown to be lower against hospitalization and severe COVID-19 due to Omicron BA.1, especially in adults, the current study demonstrates that for children and adolescents, BNT162b2 and CoronaVac continue to confer protection against COVID-19-associated hospitalization and moderate-to-severe disease.^28^ More cases of COVID-19-associated hospitalization and moderate-to-severe disease were averted by the vaccination program that began in 2021 for adolescents than vaccination that commenced during the middle of the study period in 2022 for children. VE could not be estimated for 3 doses of either vaccine because none of the 68,565 who received the third dose required hospitalization or had moderate-to-severe disease. Sensitivity analysis using 7-day lag showed roughly similar but higher VE estimates, especially for dose 1, possibly due to partial protection against severe outcomes by an early response that were classified as unvaccinated.

VE estimates against severe outcomes with Omicron BA.1 in children and adolescents have varied between studies performed in different settings. Most notably, Price and colleagues found 43% VE against COVID-19-associated hospitalizations 2-22 weeks after 2 doses of BNT162b2 in adolescents and 68% after 2 doses in children using a test-negative design in 31 pediatric hospitals in the United States.^12^ Sacco and colleagues in Italy found 41% VE against severe COVID-19 during BA.1 predominance by a population cohort analysis.^23^ Our study is unique as we analyze VE in a largely prior uninfected population, meaning our unvaccinated population will not be protected by a previous infection, for which is difficult to be completely adjusted. Additionally, hospitalization as an outcome measure varies across different settings and is imperfect as muddled by persons with incidental infection.^29^ Our data support the high VE of BNT162b2 against severe COVID-19 in adolescents and children estimated by several other groups.^15,19,21^

CoronaVac is an inactivated whole-virus vaccine that has been associated with weaker neutralization titers yet comparable or higher T cell responses than mRNA and other vaccine platforms.^10^ In this analysis, we found high protection against hospitalization and moderate-to-severe disease after 2 doses in both adolescents and children. These results are in line with analyses in Chile, Brazil and Argentina that inactivated COVID-19 vaccines elicit high protection against severe outcomes in children and adolescents.^16,17,22^ This is likely due to T cell responses elicited by the inactivated whole-virus vaccines, which are directed against all structural proteins of SARS-CoV-2 and not just the spike.^10^ We have previously shown preserved T cell responses against SARS-CoV-2 S, N and M proteins for BA.1.^30^ The current findings from this study strongly support the use of CoronaVac in children and adolescents against severe COVID-19.

Numerous VE studies must be performed as these are conducted under variable settings and the conclusions need to be interpreted in the context of all available literature, including and not solely on the current study results.^31^ Research with observational VE designs, such as the current study, can be prone to bias, which can include the possibility of residual confounding unaccounted by other factors. As Hong Kong tended to hospitalize most diagnosed cases, especially those youngest and at the very beginning of the pandemic, VE against hospitalization may be lower and closer to against infection in our dataset. However, we also evaluated VE against moderate-to-severe COVID-19, which is stringent and revealed even higher VE. Due to insufficient power, we did not evaluate VE against progression from infection to hospitalization, moderate-to-severe disease or death alone. Nevertheless, using such big datasets that already included the most essential endpoints with an adequate sample size, we were able to reach clinically pertinent conclusions. Misclassification bias is possible since some deaths occurred before hospitalization. ICD-9 coding was manually entered and could be erroneous, as detailed clinical notes were not requested to verify the ICD-9 coding. Preexisting comorbidities could not be adjusted, though they are rare in the pediatric population and not expected to influence VE estimates significantly for children and adolescents. There were few numbers of unvaccinated adolescents and thus possibly differences between the unvaccinated and vaccinated were not fully accounted. We were unable to evaluate waning of VE over time since many were recently vaccinated or received boosters, but follow-up studies have been arranged. As the HK population had very low levels of circulation of prior variants, the current findings are more applicable to parts of the world that had low past exposures. The person-time under 14 days of vaccination was categorized as the previous dose, or unvaccinated <14 days after dose 1, which can underestimate the VE of the previous dose if there was exposure around the time of vaccination. Additionally, there is the possibility that there were different social-distancing or other risk behaviors in the unvaccinated compared to the vaccinated individuals, or vice versa, that could affect the VE. However, most individuals in HK maintained strict social-distancing practices, especially during the BA.2 wave regardless of vaccination status, and masking was mandatory in public.^32^

Overall, it can be concluded that the BNT162b2 or CoronaVac vaccines confer protection against Omicron BA.2-associated severe clinical outcomes for children and adolescents, with no apparent differences observed across the two vaccine types. Public advocacy and education are essential to promote immediate vaccination in children so these individuals can benefit from timely protection against COVID-19 and more severe cases within the whole population can be averted. Further research on the impact of the third dose and durability of these vaccines in children and adolescents, and protection from vaccination in younger children ages 6 months to 2 years as the COVID-19 vaccines have become available for this age group recently, is necessary and planned.

## METHODS

### Population and study design

Since the COVID-19 pandemic, Hong Kong (HK) maintained territory-wide stringent infection control and social-distancing policies that curbed SARS-CoV-2 infection to only 13,000 cases for the 7.4-million HK population prior to its first major COVID-19 outbreak in January 2022 due to the Omicron BA.2 variant.^33,34^ This observational study’s population consisted of these Hong Kong residents aged 5-18 years for BNT162b2 and 3-18 years for CoronaVac that had very low exposures to prior SARS-CoV-2 variants. We assessed vaccine effectiveness (VE) of BNT162b2 and CoronaVac by utilizing an ecological study design. The study was approved by the Institutional Review Board of the University of Hong Kong (UW 22-217). Informed consent was not required.

### COVID-19 Vaccination Programme and vaccine coverage in Hong Kong

The emergency use of BNT162b2 and CoronaVac became authorized stepwise, first for adolescents on 21 Mar 2021 and then children on 21 Jan 2022 (Figure S1). During 16 Mar 2021 to 19 Apr 2022, COVID-19 vaccination in the pediatric population consisted of the adult formulations of BNT162b2 at the full dose of 30 micrograms for aged 12-18 years old, 10 micrograms for aged 5-11 years and 600 SU of CoronaVac for aged 3-18 years. We extracted daily infection surveillance data and vaccine coverage statistics of the whole pediatric population from DH, which included age, sex, dates of inoculation, vaccine type and dose.

### COVID-19-associated hospitalization and moderate-to-severe disease

COVID-19 has been a statutory notifiable disease in HK. The public medical system accounts for 85% of all hospitalizations and >90% for hospitalized COVID-19 in HK due to limited resources and negative pressure airborne infection isolation rooms in the private sector.^35-37^ All public hospitals in HK are managed by the Hospital Authority (HA) and utilize the Clinical Management System for electronic medical documentation, including ICD-9 coding. During January 2022 to April 2022, all cases of COVID-19-associated hospitalization and moderate-to-severe disease were confirmed by positive laboratory results using the immunofluorescence assay and/or polymerase chain reaction (PCR) on respiratory tract and/or stool specimens. In-hospital fatalities and intensive care unit (ICU) admissions associated with COVID-19 were recorded. Anonymized data on COVID-19-associated hospitalization, mortality, ICU admissions and ICD-9 numbers corresponding to the principal and secondary diagnoses, as well as procedural codes, were obtained from HA. Death, ICU admission, mechanical ventilation, respiratory therapy, pneumonia, croup (laryngotracheobronchitis) and encephalitis/encephalopathy were considered as severe COVID-19. Moderate COVID-19 comprised of febrile seizures, intestinal infection, diarrhea, nausea and vomiting since patients hospitalized for these presentations in our region commonly required intravenous fluid. Information on comorbidities pertinent to the risk of severe pediatric COVID-19 were also obtained, and the associated ICD-9 codes used are listed in Table S4.

### VE estimation and statistical analysis

The study periods for the VE estimation were 21 Jan 2022 to 19 Apr 2022 for children aged 3-11 years (vaccination began 21 Jan 2022 for this age group) and 1 Jan 2022 to 19 Apr 2022 for adolescents aged 12-18 years (vaccination began 16 Mar 2021 for this age group), with the outcome measures of COVID-19-associated hospitalization and moderate-to-severe disease during the Omicron BA.2 wave between January and April 2022 in HK. We retrieved population-level statistics from DH on the daily counts for those who received 1, 2 or 3 doses of either BNT162b2 or CoronaVac. The vaccine coverage statistics were available for the age ranges of 3-11 and 12-19, which we used to estimate the coverage in children and adolescents, respectively. The numbers of unvaccinated individuals were calculated by deducting the number of vaccinated individuals from the respective age groups in the whole population data determined by the Census and Statistics Department of the HK Government at the end of 2021.^35^ The daily counts in each vaccination category of the whole population’s uptake were estimated on the assumption that the same vaccine type was maintained for each dose, in accordance with recommendations by the HK Vaccination Programme. For hospitalized cases, DH supplied vaccination records and dates of reported infection that were linked to each patient with pseudonyms generated by DH and HA for privacy. The HA dataset contained demographic information, admission months, length of stay, clinical outcomes (death, ICU admission and hospital discharge) and ICD-9 diagnostic and procedural codes. The ICD-9 code of 519.0 (8) was used for COVID-19 (Table S4). Hospitalized patients who received heterologous prime-boost or vaccine types other than BNT162b2 and CoronaVac were excluded.

Vaccination status was categorized with a 14-day lag, the usual time period to acquire sufficient immune response against severe outcomes. Cases were stratified by ages of 3-11 years and 12-18 years. We derived the incidence rates in each vaccination category, adjusted by sex and calendar day of vaccination for each day during the specified study period, and estimated the incidence rate ratios (IRR) against the unvaccinated reference group using negative binomial regression for the daily counts of the COVID-19-associated hospitalization and moderate-to-severe disease by the date of the reported infection. The logarithm of persons-at-risk was used as the offset variable. VE was calculated by (1-IRR)×100%. To explore the impact of vaccination on the expected reduction of COVID-19-associated hospitalization and moderate-to-severe disease cases based on the VE estimates, the expected number of cases in the absence of vaccination was calculated by case_0_ + case_BNT1_/(1-VE_BNT1_) + case_BNT2_/(1-VE_BNT2_) + case_CoV1_/(1-VE_CoV1_) + case_CoV2_/(1-VE_CoV2_), in which case_0/BNT/CoV1/2_ represented the numbers of observed (actual) unvaccinated cases or cases with doses 1 or 2 of BNT162b2 or CoronaVac and VE_BNT/CoV1/2_ was the VE of doses 1 or 2 of BNT162b2 or CoronaVac, with 1-VE as the IRR.^38^ We estimated the two-sided 95%CI from the pooled IRR across the vaccine doses and types in children and adolescents using the Mantel-Haenszel method. The numbers of cases averted were derived by subtracting the observed numbers of cases from expected numbers of cases. Since there were no cases observed after dose 3, cases averted for this dose could not be estimated.

Sensitivity analyses with adjustment for calendar week instead of calendar day and 7-day instead of 14-day lag were performed. Additionally, estimation of cases averted by 2 doses of vaccination that included daily population at risk and vaccination numbers was performed by calculating the daily rate differences between the non-fully vaccinated group (unvaccinated+1 dose of BNT or CoronaVac) and vaccinated group (2 doses or more of either vaccines), with 95% CI.^39^ The daily rates were then multiplied by the numbers of the population at risk of the vaccinated group. The total numbers of estimated averted cases were calculated by summing up the daily estimated averted cases within the whole study period. There was no sample size estimation since this was a whole population study. The statistical analyses were performed with R (Version 4.0.3).

## Data Availability

Individual-level data are unable to be shared due to third party use restrictions. The aggregate dataset is appended in the source data file, and the R code is available at https://github.com/kmanyip/BA2-VE-hosp-and-mod-sev.

## ACKNOWLEDGEMENTS

We are grateful to the Centre for Health Protection, Department of Health and the Hospital Authority, both of the Hong Kong Government for providing these pertinent, pseudonymized datasets. We thank Dr Minal K. Patel from the Department of Immunization, Vaccines, and Biologicals, World Health Organization, Geneva, Switzerland for her critical review, comments and suggestions during the study planning and drafting of this manuscript.

## STATEMENT OF CONTRIBUTION

Y.L. Lau conceptualized the study. W.H.S. Wong, Y.L. Lau, J.S. Rosa Duque and D. Leung designed the study. Y.L. Lau led the acquisition of data and funding. Y.L. Lau and W.H.S. Wong supervised the project. W.H.S. Wong provided software support. W.H.S. Wong, K.M. Yip, H.K. So, J.S. Rosa Duque, D. Leung and Y.L. Lau had unrestricted access to all data. K.M. Yip, H.K. So and W.H.S. Wong curated the data and estimated the VE. K.M. Yip replicated the W.H.S. Wong oversaw the statistical analyses. J.S. Rosa Duque, D. Leung, K.M. Yip and D.H.L. Lee visualized the data. J.S. Rosa Duque and D. Leung wrote the first draft of the manuscript, which was supervised, reviewed and edited by Y.L. Lau, with input from W.H.S. Wong. All authors reviewed, approved and agreed to take full responsibility of the content of the manuscript, the accuracy of its data and fidelity of the statistical analysis.

## SUPPLEMENTAL TABLES

**Table S1.** Frequencies of comorbidities or ICD-9 code listed as the primary diagnosis for the hospitalized children (aged 3-11 years, 21 Jan 2022 to 19 Apr 2022) and adolescents (aged 12-18 years, 1 Jan 2022 to 19 Apr 2022).

| Diagnoses | ICD-9 codes | % |
| --- | --- | --- |
| **Comorbidities** |  |  |
| *No comorbidity* | N/A | 90.6% |
| *Other metabolic disorders and immunity disorders* | 270-279 | 3.4% |
| *Cardiovascular conditions* | 390-459 | 2.5% |
| *Any malignancy* | 140-239 | 1.6% |
| *Congenital anomalies* | 740-757, 759 | 0.9% |
| *Epilepsy* | 345 | 0.9% |
| *Diabetes mellitus* | 250 | 0.6% |
| *COPD and allied conditions (including asthma)* | 490-496 | 0.6% |
| *Obesity* | 278.0 | 0.4% |
| *Chromosomal anomalies* | 758 | 0.4% |
| *Hereditary and degenerative diseases of the central nervous system* | 330-337 | 0.3% |
| *Muscular dystrophies and other myopathies* | 359.0-359.2 | 0% |
| **Listed as the primary diagnosis** |  |  |
| *COVID-19* | 519.0 (8) | 87.1% |
| *Febrile seizures/seizures with fever* | 780.31, 780.32 | 11.5% |
| *Congenital anomalies* | 740-757, 759 | 0.7% |
| *Chromosomal anomalies* | 758 | 0.6% |
| *Diarrhea* | 787.91 | 0.1% |
| ICD-9, the International Classification of Diseases 9th Revision; N/A, not applicable |  |  |

**Table S2.** Vaccine effectiveness against COVID-19-associated hospitalization and moderate-to-severe disease with adjustment by calendar weeks (instead of days) and 14-day lag in children (aged 3-11 years, 21 Jan 2022 to 19 Apr 2022) and adolescents (aged 12-18 years, 1 Jan 2022 to 19 Apr 2022).

|  | Numbers of cases | Mean days elapsed since the last dose (SD) | VE estimate | 95%CI | p-value |
| --- | --- | --- | --- | --- | --- |
| **VE against hospitalization** |  |  |  |  |  |
| **Children (aged 3-11 years)** |  |  |  |  |  |
| *BNT162b2, 1 dose* | 12 | 27.4 (14.7) | 65.3% | 37.6 – 82.3 | 0.0009 |
| *CoronaVac, 1 dose* | 127 | 23.8 (7.5) | 13.0% | -14.7 – 34.2 | 0.3389 |
| *CoronaVac, 2 doses* | 4 | 23.3 (10.7) | 86.1% | 65.4 – 95.8 | 0.0001 |
| *Unvaccinated* | 967 | *ref* | *ref* | *ref* | *ref* |
| **Adolescents (aged 12-18 years)** |  |  |  |  |  |
| *BNT162b2, 1 dose* | 114 | 132.2 (54.1) | 60.2% | 47.0 – 70.1 | <0.0001 |
| *CoronaVac, 1 dose* | 22 | 34.7 (16.0) | 30.8% | -9.65 – 58.2 | 0.1317 |
| *BNT162b2, 2 doses* | 165 | 171.5 (51.2) | 82.4% | 77.0 – 86.5 | <0.0001 |
| *CoronaVac, 2 doses* | 5 | 47.4 (19.6) | 90.7% | 79.1 – 96.7 | <0.0001 |
| *Unvaccinated* | 149 | *ref* | *ref* | *ref* | *ref* |
| **VE against moderate-to-severe disease** |  |  |  |  |  |
| **Children and adolescents (aged 3-18 years)** |  |  |  |  |  |
| *BNT162b2, 1 dose* | 8 | 126.1 (54.0) | 84.6% | 69.7 – 93.2 | <0.0001 |
| *CoronaVac, 1 dose* | 33 | 22.2 (5.7) | 33.5% | -5.0 – 58.7 | 0.0883 |
| *BNT162b2, 2 doses* | 8 | 185.6 (49.4) | 93.1% | 86.4 – 97.0 | <0.0001 |
| *CoronaVac, 2 doses* | 1 | 39.0 (0.0) | 95.8% | 80.6 – 99.8 | 0.0017 |
| *Unvaccinated* | 178 | *ref* | *ref* | *ref* | *ref* |
| SD, standard deviation; VE, vaccine effectiveness; CI, confidence interval; ref, reference |  |  |  |  |  |

**Table S3.** Vaccine effectiveness against COVID-19-associated hospitalization and moderate-to-severe disease with adjustment by calendar days and 7-day (instead of 14-day) lag in children (aged 3-11 years, 21 Jan 2022 to 19 Apr 2022) and adolescents (aged 12-18 years, 1 Jan 2022 to 19 Apr 2022).

|  | Numbers of cases | Mean days elapsed since the last dose (SD) | VE estimate | 95%CI | p-value |
| --- | --- | --- | --- | --- | --- |
| **VE against hospitalization** |  |  |  |  |  |
| **Children (aged 3-11 years)** |  |  |  |  |  |
| *BNT162b2, 1 dose* | 12 | 27.4 (14.7) | 82.6% | 67.0 – 91.5 | <0.0001 |
| *CoronaVac, 1 dose* | 123 | 23.8 (7.5) | 45.4% | 23.3 – 61.2 | 0.0005 |
| *CoronaVac, 2 doses* | 8 | 23.3 (10.7) | 89.1% | 76.5 – 95.5 | <0.0001 |
| *Unvaccinated* | 968 | *ref* | *ref* | *ref* | *ref* |
| **Adolescents (aged 12-18 years)** |  |  |  |  |  |
| *BNT162b2, 1 dose* | 112 | 132.2 (54.1) | 62.9% | 48.3 – 73.5 | <0.0001 |
| *CoronaVac, 1 dose* | 18 | 34.7 (16.0) | 63.6% | 38.9 – 79.4 | 0.0003 |
| *BNT162b2, 2 doses* | 167 | 171.5 (51.2) | 79.9% | 72.5 – 85.4 | <0.0001 |
| *CoronaVac, 2 doses* | 9 | 47.4 (19.6) | 90.3% | 81.2 – 95.6 | <0.0001 |
| *Unvaccinated* | 149 | *ref* | *ref* | *ref* | *ref* |
| **VE against moderate-to-severe disease** |  |  |  |  |  |
| **Children and adolescents (aged 3-18 years)** |  |  |  |  |  |
| *BNT162b2, 1 dose* | 8 | 126.1 (54.0) | 87.9% | 76.2 – 94.7 | <0.0001 |
| *CoronaVac, 1 dose* | 33 | 22.2 (5.7) | 49.1% | 20.3 – 68.3 | 0.0043 |
| *BNT162b2, 2 doses* | 8 | 185.6 (49.4) | 93.9% | 88.0 – 97.3 | <0.0001 |
| *CoronaVac, 2 doses* | 1 | 39.0 (0.0) | 97.6% | 89.8 – 99.9 | 0.0002 |
| *Unvaccinated* | 178 | *ref* | *ref* | *ref* | *ref* |
| SD, standard deviation; VE, vaccine effectiveness; CI, confidence interval; ref, reference |  |  |  |  |  |

**Table S4.** Enumeration of the International Classification of Diseases (ICD-9) codes and other data as moderate to severe diseases and comorbidities.

| Clinical diagnoses, outcomes, procedures or conditions | ICD-9 codes |
| --- | --- |
| **Severe disease (diagnoses, outcomes or procedures)** |  |
| *Death* | (see note below)* |
| *Intensive care unit (ICU) admission* | (see note below)* |
| *Mechanical ventilation* | 96.7 |
| *Respiratory therapy* | 93.90, 93.93, 93.96, 93.99 |
| *Pneumonia* | 486.0 |
| *Croup (laryngotracheobronchitis)* | 464.4 |
| *Encephalitis/encephalopathy* | 323.9, 348.3 |
| **Moderate disease** |  |
| *Intestinal infection due to other organism, not elsewhere classified* | 008.8 |
| *Febrile seizures/seizures with fever* | 780.31, 780.32 |
| *Nausea and vomiting* | 787.01 |
| *Diarrhea* | 787.91 |
| **Comorbidities (preexisting conditions)** |  |
| *Any malignancy* | 140-239 |
| *Diabetes mellitus* | 250 |
| *Obesity* | 278.0 |
| *Other metabolic disorders and immunity disorders* | 270-279 |
| *Hereditary and degenerative diseases of the central nervous system* | 330-337 |
| *Epilepsy* | 345 |
| *Muscular dystrophies and other myopathies* | 359.0-359.2 |
| *Cardiovascular conditions* | 390-459 |
| *COPD and allied conditions (including asthma)* | 490-496 |
| *Congenital anomalies* | 740-757, 759 |
| *Chromosomal anomalies* | 758 |
| ICD-9, the International Classification of Diseases 9th Revision; *, data were provided not as ICD-9 coding |  |

## SUPPLEMENTAL FIGURES AND LEGENDS

**Figure S1.**
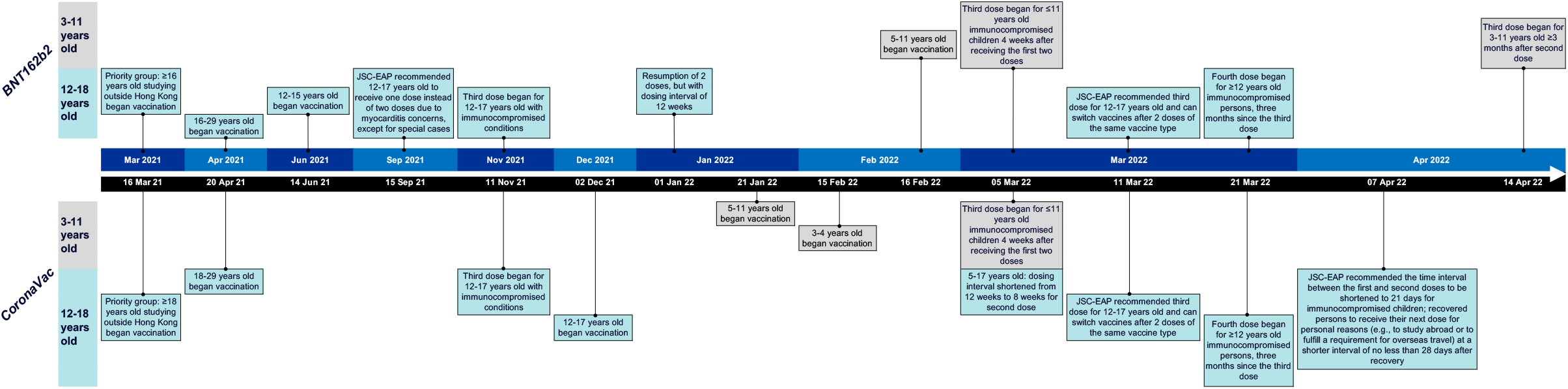
Timeline of commencement and updates on the COVID-19 vaccination scheme in Hong Kong for children aged 3-11 years and adolescents aged 12-18 years from 16 Mar 2021 to 19 Apr 2022. For adolescents aged 12-18 years, BNT162b2 commenced on 16 Mar 2021 but was allocated in sequence based on priority and age groups. The priority group consisted of those aged ≥16 years studying outside of Hong Kong, followed by other healthy individuals aged ≥16 years. Updates on approval for age reduction led to coverage down to 12 years of age, so that by December 2021, all adolescents aged 12-18 years could receive BNT162b2 or CoronaVac. Relative to adolescents, COVID-19 vaccines were approved later for children aged 3-11 years, first of which was CoronaVac in January 2022 and then BNT162b2 in February 2022. Rollout of the third dose of the BNT162b2 and CoronaVac vaccines for all healthy adolescents and children began on 11 Mar 2022 and 14 Apr 2022, respectively. JSC-EAP, the Scientific Committee on Vaccine Preventable Diseases and the Scientific Committee on Emerging and Zoonotic Diseases (JSC) under the Centre for Health Protection of the Department of Health and the Chief Executive’s expert advisory panel (EAP).

**Figure S2.**
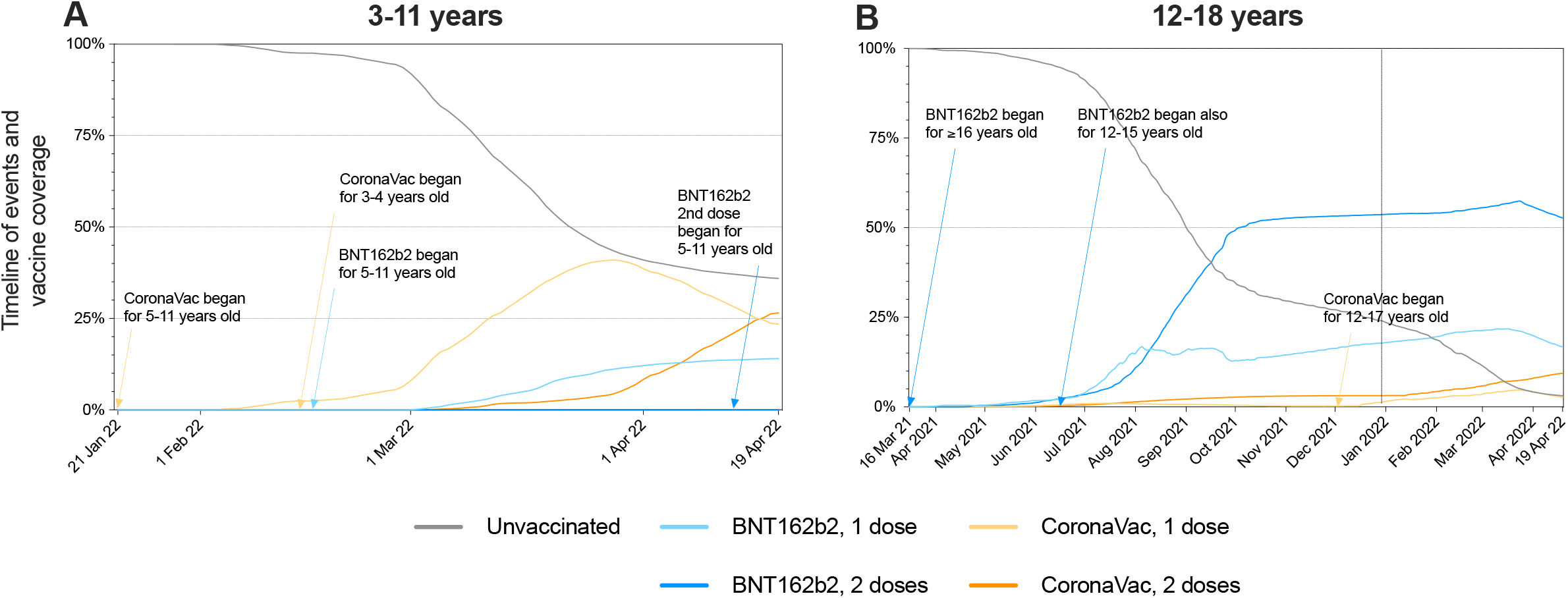
Daily counts of COVID-19 vaccinations in Hong Kong for children (commenced on 21 Jan 2022 until the end of study period of 19 Apr 2022) and adolescents (commenced on 16 Mar 2021 until the end of study period of 19 Apr 2022) (A) For children, essentially all were unvaccinated at start of the study period of 21 Jan 2022, the date of commencement of inoculation, and the numbers of vaccination rose gradually during the study period until 19 Apr 2022. (B) For adolescents, vaccination with BNT162b2 began since 16 Mar 2021, and by 1 Jan 2022, >50% had received 2 doses of BNT162b2, while CoronaVac commenced just 1 month prior (2 Dec 2021) for healthy adolescents, and <25% were unvaccinated.

**Figure S3.**
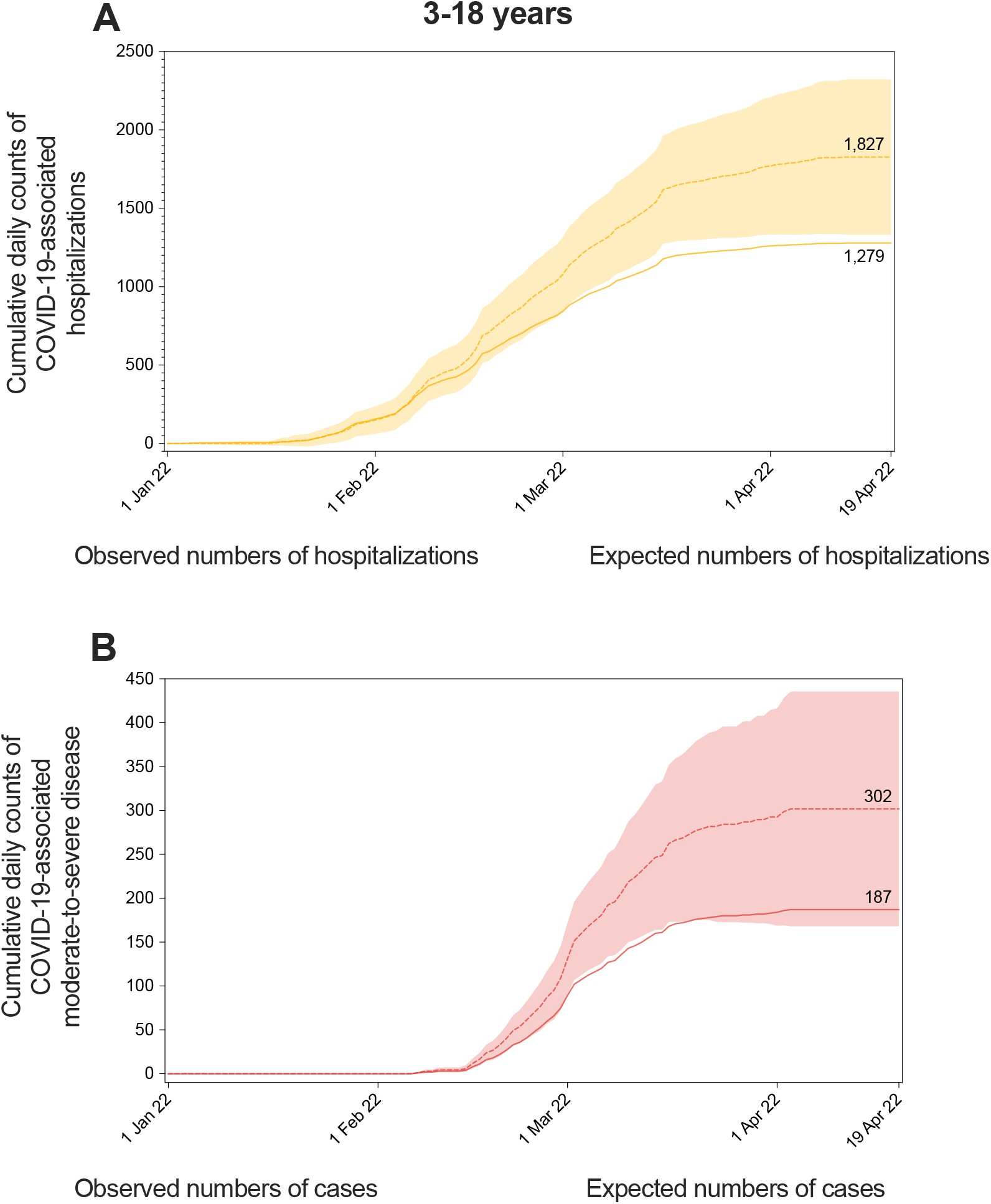
Cumulative SARS-CoV-2 outcomes over time comparing observed cases with territory-wide vaccination and predicted cases without vaccination by outcome. (A) COVID-19-associated hospitalizations, cases averted: 548 (95%CI 53-1044). (B) COVID-19-associated moderate-to-severe disease, cases averted: 115 (95%CI -19-249).

## REFERENCES

1. Tso, W.W., Kwan, M.Y., Wang, Y.L., Leung, L.K., Leung, D., Chua, G.T., Ip, P., Fong, D.Y., Wong, W.H., Chan, S.H., et al. (2022). Severity of SARS-CoV-2 Omicron BA. 2 infection in unvaccinated hospitalized children: Comparison to influenza and parainfluenza infections. Emerging Microbes & Infections. https://doi.org/10.1080/22221751.2022.2093135.

2. Martin, B., DeWitt, P.E., Russell, S., Sanchez-Pinto, L.N., Haendel, M.A., Moffitt, R., and Bennett, T.D. (2022). Acute Upper Airway Disease in Children With the Omicron (B.1.1.529) Variant of SARS-CoV-2-A Report From the US National COVID Cohort Collaborative. JAMA Pediatr 176, 819–821. 10.1001/jamapediatrics.2022.1110.

3. American Academy of Pediatrics (2022). Children and COVID-19: State Data Report, A joint report from the American Academy of Pediatrics and the Children’s Hospital Association, Version 8/18/22..

4. United Nations Children’s Fund (UNICEF) (2022). COVID-19 confirmed cases and deaths. https://data.unicef.org/resources/covid-19-confirmed-cases-and-deaths-dashboard/.

5. Mallapaty, S., Callaway, E., Kozlov, M., Ledford, H., Pickrell, J., and Van Noorden, R. (2021). How Covid vaccines shaped 2021 - in eight powerful charts. Nature 600 (23/30), 580–583.

6. Leung, D., Duque, J.R., Yip, K.M., So, H.K., Wong, W., and Lau, Y.L. (2022). Effectiveness of BNT162b2 and CoronaVac in children and adolescents against SARS-CoV-2 infection during Omicron BA.2 wave in Hong Kong. Res Sq. 10.21203/rs.3.rs-1856540/v1.

7. Frenck, R.W., Jr., Klein, N.P., Kitchin, N., Gurtman, A., Absalon, J., Lockhart, S., Perez, J.L., Walter, E.B., Senders, S., Bailey, R., et al. (2021). Safety, Immunogenicity, and Efficacy of the BNT162b2 Covid-19 Vaccine in Adolescents. N Engl J Med 385, 239–250. 10.1056/NEJMoa2107456.

8. Walter, E.B., Talaat, K.R., Sabharwal, C., Gurtman, A., Lockhart, S., Paulsen, G.C., Barnett, E.D., Muñoz, F.M., Maldonado, Y., Pahud, B.A., et al. (2022). Evaluation of the BNT162b2 Covid-19 Vaccine in Children 5 to 11 Years of Age. N Engl J Med 386, 35–46. 10.1056/NEJMoa2116298.

9. Han, B., Song, Y., Li, C., Yang, W., Ma, Q., Jiang, Z., Li, M., Lian, X., Jiao, W., Wang, L., et al. (2021). Safety, tolerability, and immunogenicity of an inactivated SARS-CoV-2 vaccine (CoronaVac) in healthy children and adolescents: a double-blind, randomised, controlled, phase 1/2 clinical trial. The Lancet Infectious Diseases 21, 1645–1653. 10.1016/s1473-3099(21)00319-4.

10. Rosa Duque, J., Wang, X., Leung, D., Cheng, S., Cohen, C., Mu, X., Hachim, A., Zhang, Y., Chan, S.-M., Chaothai, S., et al. (2022). Immunogenicity and reactogenicity of SARS-CoV-2 vaccines BNT162b2 and CoronaVac in healthy adolescents. Nat Commun 13, 3700. https://doi.org/10.1038/s41467-022-31485-z.

11. Fleming-Dutra, K.E., Britton, A., Shang, N., Derado, G., Link-Gelles, R., Accorsi, E.K., Smith, Z.R., Miller, J., Verani, J.R., and Schrag, S.J. (2022). Association of Prior BNT162b2 COVID-19 Vaccination With Symptomatic SARS-CoV-2 Infection in Children and Adolescents During Omicron Predominance. JAMA 327, 2210–2219. 10.1001/jama.2022.7493.

12. Price, A.M., Olson, S.M., Newhams, M.M., Halasa, N.B., Boom, J.A., Sahni, L.C., Pannaraj, P.S., Irby, K., Bline, K.E., Maddux, A.B., et al. (2022). BNT162b2 Protection against the Omicron Variant in Children and Adolescents. N Engl J Med 386, 1899–1909. 10.1056/NEJMoa2202826.

13. Veneti, L., Berild, J.D., Watle, S.V., Starrfelt, J., Greve-Isdahl, M., Langlete, P., Bøås, H., Bragstad, K., Hungnes, O., and Meijerink, H. (2022). Vaccine effectiveness with BNT162b2 (Comirnaty, Pfizer-BioNTech) vaccine against reported SARS-CoV-2 Delta and Omicron infection among adolescents, Norway, August 2021 to January 2022. medRxiv, 2022.2003.2024.22272854. 10.1101/2022.03.24.22272854.

14. Amir, O., Goldberg, Y., Mandel, M., Bar-On, Y.M., Bodenheimer, O., Freedman, L., Ash, N., Alroy-Preis, S., Huppert, A., and Milo, R. (2022). Initial protection against Omicron in children and adolescents by BNT162b2. medRxiv, 2022.2005.2022.22275323. 10.1101/2022.05.22.22275323.

15. Tan, S.H.X., Cook, A.R., Heng, D., Ong, B., Lye, D.C., and Tan, K.B. (2022). Effectiveness of BNT162b2 Vaccine against Omicron in Children 5 to 11 Years of Age. N Engl J Med 387, 525–532. 10.1056/NEJMoa2203209.

16. Jara, A., Undurraga, E.A., Zubizarreta, J.R., Gonzalez, C., Acevedo, J., Pizarro, A., Vergara, V., Soto-Marchant, M., Gilabert, R., Flores, J.C., et al. (2022). Effectiveness of CoronaVac in children 3-5 years of age during the SARS-CoV-2 Omicron outbreak in Chile. Nat Med. 10.1038/s41591-022-01874-4.

17. Florentino, P.T.V., Alves, F.J.O., Cerqueira-Silva, T., Oliveira, V.A., Júnior, J.B.S., Jantsch, A.G., Penna, G.O., Boaventura, V., Werneck, G.L., Rodrigues, L.C., et al. (2022). Vaccine effectiveness of CoronaVac against COVID-19 among children in Brazil during the Omicron period. Nat Commun 13, 4756. 10.1038/s41467-022-32524-5.

18. Klein, N.P., Stockwell, M.S., Demarco, M., Gaglani, M., Kharbanda, A.B., Irving, S.A., Rao, S., Grannis, S.J., Dascomb, K., Murthy, K., et al. (2022). Effectiveness of COVID-19 Pfizer-BioNTech BNT162b2 mRNA Vaccination in Preventing COVID-19–Associated Emergency Department and Urgent Care Encounters and Hospitalizations Among Nonimmunocompromised Children and Adolescents Aged 5–17 Years — VISION Network, 10 States, April 2021–January 2022. Morbidity and Mortality Weekly Report 71, 352–358.

19. Fowlkes, A.L., Yoon, S.K., Lutrick, K., Gwynn, L., Burns, J., Grant, L., Phillips, A.L., Ellingson, K., Ferraris, M.V., LeClair, L.B., et al. (2022). Effectiveness of 2-Dose BNT162b2 (Pfizer BioNTech) mRNA Vaccine in Preventing SARS-CoV-2 Infection Among Children Aged 5–11 Years and Adolescents Aged 12–15 Years — PROTECT Cohort, July 2021–February 2022. Morbidity and Mortality Weekly Report 71, 422–428.

20. Cohen-Stavi, C.J., Magen, O., Barda, N., Yaron, S., Peretz, A., Netzer, D., Giaquinto, C., Judd, A., Leibovici, L., Hernán, M.A., et al. (2022). BNT162b2 Vaccine Effectiveness against Omicron in Children 5 to 11 Years of Age. N Engl J Med. 10.1056/NEJMoa2205011.

21. Buchan, S.A., Nguyen, L., Wilson, S.E., Kitchen, S.A., and Kwong, J.C. (2022). Vaccine Effectiveness of BNT162b2 Against Delta and Omicron Variants in Adolescents. Pediatrics. 10.1542/peds.2022-057634.

22. Gonzalez, S., Olszevicki, S., Gaiano, A., Baino, A.N.V., Regairaz, L., Salazar, M., Pesci, S., Marin, L., Martinez, V.V.G., Varela, T., et al. (2022). Effectiveness of BBIBP-CorV, BNT162b2 and mRNA-1273 vaccines against hospitalisations among children and adolescents during the Omicron outbreak in Argentina: A retrospective cohort study. Lancet Reg Health Am 13, 100316. 10.1016/j.lana.2022.100316.

23. Sacco, C., Del Manso, M., Mateo-Urdiales, A., Rota, M.C., Petrone, D., Riccardo, F., Bella, A., Siddu, A., Battilomo, S., Proietti, V., et al. (2022). Effectiveness of BNT162b2 vaccine against SARS-CoV-2 infection and severe COVID-19 in children aged 5-11 years in Italy: a retrospective analysis of January-April, 2022. The Lancet 400, 97–103. 10.1016/S0140-6736(22)01185-0.

24. Cheng, S.M., Mok, C.K.P., Chan, K.C., Ng, S.S., Lam, B.H., Luk, L.L., Ko, F.W., Chen, C., Yiu, K., Li, J.K., et al. (2022). SARS-CoV-2 Omicron variant BA.2 neutralisation in sera of people with Comirnaty or CoronaVac vaccination, infection or breakthrough infection, Hong Kong, 2020 to 2022. Euro Surveill 27. 10.2807/1560-7917.ES.2022.27.18.2200178.

25. Mykytyn, A.Z., Rissmann, M., Kok, A., Rosu, M.E., Schipper, D., Breugem, T.I., Doel, P.B.v.d., Chandler, F., Bestebroer, T., Wit, M.d., et al. (2022). Antigenic cartography of SARS-CoV-2 reveals that Omicron BA.1 and BA.2 are antigenically distinct. Sci Immunol, abq4450.

26. Cowling, B.J., Ali, S.T., Ng, T.W.Y., Tsang, T.K., Li, J.C.M., Fong, M.W., Liao, Q., Kwan, M.Y.W., Lee, S.L., Chiu, S.S., et al. (2020). Impact assessment of non-pharmaceutical interventions against coronavirus disease 2019 and influenza in Hong Kong: an observational study. The Lancet Public Health 5, e279–e288. 10.1016/s2468-2667(20)30090-6.

27. Ali, S.T., Wang, L., Lau, E.H.Y., Xu, X.-K., Du, Z., Wu, Y., Leung, G.M., and Cowling, B.J. (2020). Serial interval of SARS-CoV-2 was shortened over time by nonpharmaceutical interventions. Science 369, 1106–1109.

28. Higdon, M.M., Baidya, A., Walter, K.K., Patel, M.K., Issa, H., Espié, E., Feikin, D.R., and Knoll, M.D. (2022). Duration of effectiveness of vaccination against COVID-19 caused by the omicron variant. The Lancet Infectious Diseases. 10.1016/s1473-3099(22)00409-1.

29. Feikin, D.R., Abu-Raddad, L.J., Andrews, N., Davies, M.A., Higdon, M.M., Orenstein, W.A., and Patel, M.K. (2022). Assessing vaccine effectiveness against severe COVID-19 disease caused by omicron variant. Report from a meeting of the World Health Organization. Vaccine 40, 3516–3527. 10.1016/j.vaccine.2022.04.069.

30. Leung, D., Cohen, C.A., Mu, X., Rosa Duque, J., Cheng, S.M., Wang, X., Wang, M., Zhang, W., Zhang, Y., Tam, I.Y., et al. (2022). Immunogenicity Against Wild-Type and Omicron SARS-CoV-2 After a Third Dose of Inactivated COVID-19 Vaccine in Healthy Adolescents. SSRN: Cell Press Sneak Peek. https://ssrn.com/abstract=4115862.

31. Feikin, D.R., Higdon, M.M., Abu-Raddad, L.J., Andrews, N., Araos, R., Goldberg, Y., Groome, M.J., Huppert, A., O’Brien, K.L., Smith, P.G., et al. (2022). Duration of effectiveness of vaccines against SARS-CoV-2 infection and COVID-19 disease: results of a systematic review and meta-regression. The Lancet 399, 924–944. 10.1016/s0140-6736(22)00152-0.

32. Cheng, V.C., Wong, S.C., Chuang, V.W., So, S.Y., Chen, J.H., Sridhar, S., To, K.K., Chan, J.F., Hung, I.F., Ho, P.L., and Yuen, K.Y. (2020). The role of community-wide wearing of face mask for control of coronavirus disease 2019 (COVID-19) epidemic due to SARS-CoV-2. J Infect 81, 107–114. 10.1016/j.jinf.2020.04.024.

33. HK Centre for Health Protection (2022). Statistics. https://www.chp.gov.hk/en/static/24012.html#. 2022.

34. Chiu, S.S., Cowling, B.J., Peiris, J.S.M., Chan, E.L.Y., Wong, W.H.S., and Lee, K.P. (2022). Effects of Nonpharmaceutical COVID-19 Interventions on Pediatric Hospitalizations for Other Respiratory Virus Infections, Hong Kong. Emerg Infect Dis 28, 62–68. 10.3201/eid2801.211099.

35. Census and Statistics Department of the HKSAR (2021). Hong Kong Annual Digest of Statistics (2021 Edition). https://www.censtatd.gov.hk/en/data/stat_report/product/B1010003/att/B10100032021AN21B0100.pdf.

36. news.gov.hk (2022). HK reports 6.4k COVID-19 cases. https://www.news.gov.hk/eng/2022/08/20220823/20220823_171411_923.html.

37. The Government of the Hong Kong Special Administrative Region (2022). Utilisation of private hospital beds by patients from the Hospital Authority. https://gia.info.gov.hk/general/202208/24/P2022082400919_400181_1_1661351397143.pdf.

38. UK Health Security Agency (2022). COVID-19 vaccine surveillance report: Week 19. https://assets.publishing.service.gov.uk/government/uploads/system/uploads/attachment_data/file/1075115/COVID-19_vaccine_surveillance_report_12_May_2022_week_19.pdf.

39. Haas, E.J., McLaughlin, J.M., Khan, F., Angulo, F.J., Anis, E., Lipsitch, M., Singer, S.R., Mircus, G., Brooks, N., Smaja, M., et al. (2022). Infections, hospitalisations, and deaths averted via a nationwide vaccination campaign using the Pfizer–BioNTech BNT162b2 mRNA COVID-19 vaccine in Israel: a retrospective surveillance study. The Lancet Infectious Diseases 22, 357–366. 10.1016/s1473-3099(21)00566-1.

